# Association between gait speed decline and EEG abnormalities in a cohort of active older adults living in the community

**DOI:** 10.1101/2020.05.03.20089540

**Authors:** Daysi García-Agustin, Rosa Ma. Morgade-Fonte, María A Bobes, Lídice Galán-García, Valia Rodríguez-Rodríguez

**Affiliations:** Cuban Centre for Longevity, Ageing and Health Studies, Cuba; Cuban Neuroscience Centre, Cuba; Aston University, United Kingdom

**Author notes:** Corresponding author: Valia Rodríguez-Rodríguez, School of Life and Health Sciences, Aston University, Aston Triangle, Birmingham, B4 7ET, United Kingdom. Declaration of Funding: This work was supported in part by Aston University - Global Challenge Research Fund (ID 28014) granted to V Rodríguez-Rodríguez and D García-Agustin.

**Keywords:** gait speed, community-dwelling, older adults, EEG, quantitative EEG

## Abstract

**Background:** Age-related gait speed decline is associated with pathological brain changes. Despite EEG sensitivity for detecting alteration in the brain function, its clinical use to evaluate walking disorders is rare. In this study, we examined the association of subclinical gait speed deterioration in older individuals with changes in the EEG frequency composition as a marker of mild cerebral dysfunction.

**Methods:** We studied the gait speed of 90 physically active, community-dwelling older adults in two different appointments separated by an interval of 6 years. In the last assessment, resting-state EEG was acquired in all participants. After a visual clinical report, the EEG was transformed to the frequency domain, and the energy of each frequency band was compared against normative data.

**Results:** 63 of 90 older individuals had gait speed deterioration, 87% of which also had an abnormal EEG frequency composition. EEG abnormalities were characterised by an increase in delta and theta bands power, higher than that expected for their age. We found some participants with gait speed decline that also had a decrease in alpha band power. EEG sensitivity for classifying participants with a deterioration in gait speed was 0.86, with a likelihood ratio of 2.9.

**Conclusions:** The subclinical gait speed decline found in this cohort was associated with significant changes in resting-state EEG frequency composition. The sensitivity of EEG frequency abnormalities for identifying older individuals with deterioration in gait speed supports its inclusion in the assessment pathway of older individuals at risk of developing gait disorders.

**Key Points:**

- Independent and physically active older individuals had a sub-clinical gait speed deterioration.
- Gait speed decline was associated with abnormalities in the EEG frequency composition not expected for their age.
- EEG frequency changes had a high sensitivity for classifying older adults with gait speed decline.
- Inclusion of quantitative EEG in the assessment pathway of older adults at risk of mobility disorders could serve in the identification of associated mild brain dysfunction and help in decisions related to appropriate clinical management.

## Introduction

Ageing is characterised by normal progressive changes leading to functional decline. A major risk in healthy ageing is the development of a frailty state due to a breakage in the homeostatic reserve. Disturbance in physiological homeostasis leading to frailty has widespread effects on different body organs 1–3. The musculoskeletal system is affected by such disruption, but the brain is also altered, potentially compromising the cognitive control and implementation of motor actions 4. The occurrence of any degree of brain dysfunction will necessarily affect physical performance 5,6.

Walking speed (or gait speed, GS) is the physical measure most widely used in clinical practice to evaluate the physical condition of older individuals ^7–11^. It was included as a feature of the fragility phenotype described by Fried and co-workers ^12^ that identifies older adults at risk of disability. GS has been assessed in multiple cross-sectional and longitudinal studies, not only as an indicator of the health status of older individuals but also as a predictor of adverse health conditions leading to institutionalisation, hospitalisation, and death^7,10,13,14^.

Neuroimaging techniques have proven highly valuable for identifying brain abnormalities associated with gait disorders. From these studies, it is known that gait speed decline is associated with grey matter atrophy in the prefrontal area and a higher volume of white matter hyperintensities^15,16^. Unfortunately, neuroimaging techniques are costly and may not be easy to access; this precludes their inclusion in routine assessments for the diagnosis and management of gait disorders in the geriatric clinic.

Electroencephalography (EEG) can be a low cost and accessible alternative. EEG provides valuable information on changes in brain function, with high temporal resolution. Given these advantages and its sensitivity in identifying functional patterns related to healthy and pathological ageing^17,18^, EEG and EEG-based applications have been used for research to investigate different aspects of motor control physiology^19,20^. Nevertheless, it is very rare to use EEG for clinical assessment of possible brain dysfunction associated with age-related walking disorders. In this study, we evaluated whether changes in GS measured in a cohort of active, community-dwelling older adults were associated with alterations in EEG frequency composition as a marker of mild cerebral dysfunction.

## Methods

We ran a prospective study in 2010 and 2016, involving 90 community-dwelling participants over 60 years old that regularly practised mild-to-moderate exercise in their communities. Table 1 shows the sample composition of the cohort regarding age, sex, known chronic diseases and score in the Mini-Mental State Exam. All participants provided written informed consent. The study was approved by the Ethics Committee of the Cuban Centre for Longevity, Ageing, and Health Studies.

**Table 1.** Demographic composition of the cohort included in this study.

| Characteristics | N=95 |
| --- | --- |
| Age | 64-97 (Mean=79.2, SD=7.4) |
| MMSE score | Mean=28.2, SD=2.02 |
|  | **Proportion, %** |
| Female | 87 (91.5) |
| Diabetes Mellitus | 13 (19.9) |
| Hypertension | 42 (41.6) |
| History of Stroke | 3 (3.1) |

Participants were divided into three groups according to the gait speed (GS) value measured in the 2010 and 2016 evaluation. GS was quantified as the time spent to cover 4 meters at a normal pace and was expressed in metres per second (m/sec). The composition of the groups was as follows: Group GG, 27 participants with preserved GS (i.e. >0.8 m/sec) in both 2010 and 2016 evaluations; Group GB, 32 participants with normal GS in 2010 but slower GS (<0.8 m/sec) in 2016; Group BB, 31 participants with abnormal GS (<0.8 m/sec) in both evaluations.

Besides GS assessment, an EEG recording was carried out during the 2016 appointment. EEG was obtained using a MEDICID 5-32 system (Neuronic S.A.). Nineteen Ag/AgCl electrodes were placed on the scalp according to the international 10/20 electrode placement system with the reference electrode on linked earlobes. Electrode impedance was kept below 10 kOhm. The following acquisition parameters were employed: gain of 10,000; pass-band filters between 0.3-30 Hz, 60 Hz “notch” filter, sampling interval of 5 msec, and environmental temperature of approximately 23°C. EEG was acquired for a total time of 10 min during which participants were asked to close and open their eyes at different moments to explore reactivity but also to avoid somnolence.

After a standard visual evaluation of EEG performed for clinical purposes, a quantitative analysis was carried out on the recording for each participant. Artefact-free segments of 2.56-sec duration were manually selected from eyes-closed periods by an expert electroencephalographer. About 24 epochs (~ 2 min in total) obtained from each participant were submitted to a frequency analysis using a Fast Fourier Transform (FFT). Power spectra were obtained from 0.39 to 19.11 Hz at steps of 0.39Hz. Absolute Power (AP) from the four classic frequency bands - delta, theta, alpha and beta – (here referred as broad-band spectral parameters, BBSP) were calculated and compared to the Cuban normative data of quantitative EEG (qEEG) ^21,22^.

The Z-transformed statistic was used to compare each participant’s BBSP against the normative data. This transformation expresses the distance between an individual BBSP and the average BBSP of the normal population for the participant’s age. The distance is measured in values of standard deviations (SD). If | Z | > 1.96, this means that the participant’s parameter is outside the range of the normal population with a 5% risk of error (p<0.05). Here, a participant’s BBSP was considered as abnormal if the Z value of AP for any frequency band and derivation was two or more SDs from the normal population value. The sensitivity and specificity of this classification (i.e. abnormal BBSP using |Z| > 1.96) to sort participants into the bad (BB) gate speed group were calculated. Finally, the association between GS abnormality in 2010 and in 2016, as well as between an abnormal qEEG and decline in GS, was determined using a Chi-square test.

## Results

Table 2 shows the distribution of older adults per group according to the results of the BBSP analysis. 62 of 90 participants (69%) had frequency abnormalities in the EEG. Most of these abnormalities (54/62, 87%) were present in participants with a below-threshold GS at the 2016 appointment (i.e. BB with 28/62 and GB with 26/62). EEG sensitivity and specificity to categorise participants as having a bad GS in 2016, considering the presence of qEEG abnormalities, was 0.86 and 0.70 respectively, for a likelihood ratio of 2.9, a positive predictive value of 0.87 and a negative predictive value of 0.68.

**Table 2.** Proportion of normal and abnormal qEEG studies in each gait speed group. GG: normal GS in 2010 and 2016, GB: good GS in 2010 but deteriorated in 2016, BB: gait speed deterioration in 2010 and 2016.

| Groups | Normal qEEG | Abnormal qEEG |
| --- | --- | --- |
| **GG (N=27)** | 19/27 (70.3%) | 8/27 (29.6%) |
| **GB (N=32)** | 6/32 (18.7%) | 26/32 (81.2%) |
| **BB (N=31)** | 3/31 (9.6%) | 28/31 (90.3%) |
| **Total (N=90)** | 28/90 (31.1%) | 62/90 (68.8%) |

Table 3 shows the specific types of abnormalities by frequency band found in each group. An increment in the energy of delta and theta bands was observed in groups BB and GB. Specifically, 17 of 28 individuals (61 %) in BB and 20 of 26 individuals (77%) in GB had this type of qEEG alteration. Thirteen participants in these two groups (4 in GB and 9 in BB) also had a reduction in the energy of alpha band. On the other hand, despite normal values of GS in both assessment visits, some participants (6/27, 22%) in GG also had increases in the slow frequency bands. However, no GG participant showed abnormalities in alpha power. Changes in beta band power were equally distributed among the three groups.

**Table 3.** Proportion of abnormal changes per clinical EEG frequency band in each gait speed group. The frequency band was considered abnormal if its power differed more than 1.96 SD from the population normative data for the corresponding age.

| Groups | ↑ Delta | ↑ Theta | ↓ Alpha | ↑ Beta |
| --- | --- | --- | --- | --- |
| **GG** | 2 | 4 | 0 | 2 |
| **GB** | 7 | 13 | 4 | 2 |
| **BB** | 8 | 9 | 9 | 2 |
| **Total** | 17 | 26 | 13 | 6 |

Finally, the presence of a below-threshold GS in 2016 was associated with a previous below-threshold GS in 2010 (X^2^(1) = 27.4, p<0.001). The deterioration in GS found at both assessment time points - 2010 and 2016 - was also significantly associated with qEEG abnormalities recorded in the 2016 appointment (X^2^(1) = 7.38, p<0.001; X^2^(1) = 5.76, p=0.01, respectively).

## Discussion

Our findings showed that most of the participants with a reduction in gait speed had alterations in EEG frequency composition. Specifically, slower gait speed in this older sample was associated with a higher amount of slow frequency activity than expected for their age. Interestingly, the presence of EEG slowing in the second appointment was associated with gait decline even if GS was within normal limits in the first appointment. However, since there were six years between assessments, the temporal relationship between GS deterioration and the increment in the EEG slow activity is unclear. It is possible that subclinical decline in the GS of participants with normal values in 2010 could have started soon after this assessment, and, as a consequence, the temporal evolution of the subclinical decline was similar to the other participants with manifest abnormalities in 2010.

Changes in EEG composition related to physiological ageing are characterised by the slowing of background activity, as well as a decrease in alpha power concomitant with an increase of delta and theta power^17^. However, there is still some debate regarding the extent to which slow activity is a normal pattern in healthy ageing^23^. In this study, specific frequency abnormalities were only identified if the individual frequency band deviated more than 2 SD from the normative data for the corresponding age. The sensitivity of EEG abnormalities to classify individuals as having a below-threshold gait speed was high with a good positive predictive value. Our findings seem to suggest that a significant difference in the EEG frequency composition in older individuals, when compared to a normative database, could be used as a marker of subclinical gait speed disorders. Specifically, power increase in slow frequencies and power decrease in the alpha band appears to represent a three-times greater risk for the presence of the GS disorder in this population.

Interestingly, the sample included in this study was of independent and physically active older individuals. The alteration in gait speed found in 70% of this sample represented a subclinical functional decline that suggests a higher vulnerability to minor stressors and to becoming frail. In fact, gait speed decline is one of the main features that characterise a frailty state at an older age. The ageing brain, like other organs of the body, is susceptible to the processes leading to physical frailty; therefore, we expect a degree of brain dysfunction in individuals with age-related physical decline. The association of mild brain dysfunction and gait speed decline is supported by the frequent association of neurological alterations with physical deterioration in older persons without primary neurological diseases^24^. Since EEG results from the projection onto the scalp of synchronic postsynaptic potentials generated across large cortical areas, the signal is highly sensitive to alterations in the brain function. Any disruption in neuronal communication, such as that produced by metabolic disorders or changes in underlying neural network structure, will alter the frequency composition of the EEG signal. It is not unlikely then that the EEG frequency changes associated with abnormal GS found in this study reflect the presence of subacute or chronic mild brain dysfunction in these individuals.

Furthermore, the control of gait is a process that, besides subcortical and peripheral integration of sensorimotor signals, also depends on other complex processes comprising high-level control^25^. Brain areas related to movement processing such as the supplementary motor area, premotor area and the sensorimotor cortex, interact with multi-demand regions such as the dorsal prefrontal cortex and cingulate area, to anticipate, plan, prepare, implement and readjust motor acts^25^.

During ageing, this cognitive control of movement increases^26^, possibly, as a compensatory mechanism to counteract age-related neurodegeneration^27^. Given the greater involvement of the brain in the control of the musculoskeletal system with ageing, we expect that older individuals will have changes in normal EEG patterns in relation to motor decline.

Electrophysiology is a useful tool to address questions related to the physiopathology and underlying substrate of gait deterioration^28,29^. Clinical EEG and its quantitative analysis also have the potential to provide valuable information to complement clinical evaluation for diagnosis and follow-up of frail individuals. To our knowledge, this is the first report to use quantitative EEG, as currently applied in a clinical setting, to explore gait speed deterioration in community-dwelling older adults. Our findings need to be replicated and validated in an independent sample, but they suggest that independent older individuals can develop a mild brain dysfunction associated with subclinical physical decline. If that is true, early detection could prompt early management to delay or prevent the development of a frailty state. Finally, this study supports the use and value of qEEG for the assessment of older individuals at risk of developing gait disorders as well as the incorporation of qEEG into diagnostic and rehabilitation pathways of frailty.

## Data Availability

Data will be available upon request after publication

